# The transmission of SARS-CoV-2 is likely comodulated by temperature and by relative humidity

**DOI:** 10.1101/2020.05.23.20111278

**Authors:** Kevin S. Raines, Sebastian Doniach, Gyan Bhanot

## Abstract

Inferring the impact of climate upon the transmission of SARS-CoV-2 has been confounded by variability in testing, unknown disease introduction rates, and changing weather. Here we present a data model that accounts for dynamic testing rates and variations in disease introduction rates. We apply this model to data from Colombia, whose varied and seasonless climate, central port of entry, and swift, centralized response to the Covid-19 pandemic present an opportune environment for assessing the impact of climate factors on the spread of Covid-19. We observe strong attenuation of transmission in climates with sustained daily temperatures above 30 degrees Celsius and simultaneous mean relative humidity below 78%, with outbreaks occurring at high humidity even where the temperature is high. We hypothesize that temperature and relative humidity comodulate the infectivity of SARS-CoV-2 within respiratory droplets.

## Introduction

Coronaviruses are a class of large, enveloped, single-strand RNA viruses that are widespread in animals and provoke respiratory illnesses in humans [1]. The novel coronavirus SARS-CoV-2 was identified in January 2020 as the likely causative agent of a cluster of pneumonia cases appearing in Wuhan, China throughout December 2019, making it the seventh known coronavirus to cause pathology in humans [2]. SARS-CoV-2 is associated with a respiratory illness, Covid-19, that ranges in severity from an asymptomatic infection [3], to common-cold like symptoms, to viral pneumonia, acute respiratory distress syndrome, and death [4]. While the mortality in SARS-CoV-2 appears to be lower than in SARS-CoV [4], this new virus has more effective transmission characteristics [5], including an asymptomatic infective phase [6].

From early January to March 2020, SARS-CoV-2 quickly spread around the world, causing a global pandemic [7]. While global data will certainly play a role in elucidating the epidemiology of Covid-19, we have identified three factors that confound global data: asynchronous and varied responses by local and national governments [7, 8], regional variations in disease awareness, testing protocols, and testing kit-availability leading to disparate and dynamic disease detection rates [9] and unknown variations in the rate at which infectious travelers carried the disease into local populations [10].

Varied responses by governments, from total quarantine to vague social distancing suggestions, dynamic detection rates (Fig. 1) and varying rates at which the disease was introduced into different regions each present unique challenges to the analysis of global data. Changing seasons in the hemispheres during the spread of the pandemic further confounds the analysis of weather factors.

**Fig 1.**
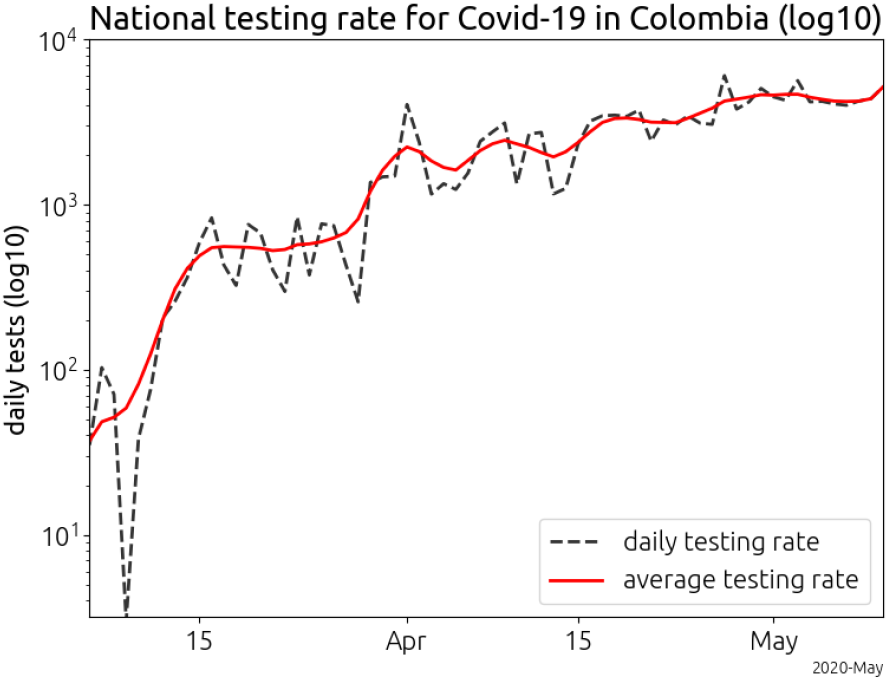
Testing rates in Colombia, March through early May. The number of daily tests for SARS-CoV-2 increased by two orders of magnitude over two months. We have identified dynamic testing rates as a key confounding factor in the analysis of Covid-19 case data.

In order to mitigate these factors, we we developed a data model and conducted a by-city analysis of data from Colombia. Colombia is uniquely suited for the study of weather factors on the transmission of SARS-CoV-2 for the following five reasons:

### 1. Climate variation

Colombia has five geographically distinct regions: The Pacific coastal region, the Caribbean coastal region, the Andean mountain region, *los llanos* (grassland plains), and the Amazon Rainforest region. The unique and seasonless climate in each of these regions alleviates the confounding role of seasons on data from the hemispheres [11].

### 2. Central port of entry

The El Dorado International airport in Bogotáis by far the largest transportation hub for international travel, with nearly seven times the number of international travelers as the next largest airport in Colombia.

### 3. Conditions favorable to rapid spread

Colombian cities have high urban population densities (Table 1), and public transportation is widely used in Colombia, with only one fifth the number of registered cars per citizen compared to the USA. For example, in Barranquilla, a coastal city in the Caribbean with hot temperatures, the average weekday commute time on public transportation is 77 minutes. The heavy use of crowded public transportation means that the measured lack of spread of the virus in this city is not the result of social distancing among the populace.

**Table 1.** Geographic and population data for the five largest cities in Colombia.

|  | population | pop. density | max. daily temp | daily humidity |
| --- | --- | --- | --- | --- |
| Bogotá | 7,387,400 | 19,475 | 20.7 | 82 |
| Medellín | 2,382,399 | 20,681 | 26.0 | 83 |
| Cali | 2,172,527 | 16,247 | 27.0 | 81 |
| Barranquilla | 1,205,284 | 14,455 | 32.3 | 68 |
| Cartagena | 876,885 | 17,053 | 30.1 | 74 |

### 4. Lack of air-conditioning

Indoor air-conditioning is rare in Colombia [12]. The citizens live in the ambient conditions of temperature and humidity of their environment. This eliminates individual specific variation in temperature and humidity as a potential confounding factor.

### 5. Swift and coordinated national response

The first Covid-19 case was confirmed in Colombia on March 6, 2020. Nineteen days later, the government implemented a national quarantine, with tight cooperation at the local level and testing orchestrated centrally through Bogotá[13]. This strong, centralized and swift action at the national level greatly simplifies the analysis because the data cleanly separates into The temperature is the maximum daily temperature and the humidity is the mean daily relative humidity. The values shown are averages taken over the month of March, 2020. The population density is the urban population density.

#### pre-quarantine and post-quarantine periods

The *transmission rate*, or the propensity of the disease to spread, must be distinguished from both total disease prevalence and the number of confirmed cases in a given population. As we show in the supplementary text, the the number confirmed cases is confounded by the detection rate and the drip rate, or the rate at which infectious travelers arrive in a city. *Dynamic* detection rates caused by increasing disease awareness and testing introduce an additive factor into the exponent of the apparent transmission rate (supporting information). Over the course of our study, the daily testing rate in Colombia increased by over two orders of magnitude (Fig. 1). Our data model accounts for variations in the drip rate and dynamic detection rates by considering confirmed case dynamics instead of total confirmed case numbers, since totals are confounded by public perception, varying testing rates, varying drip rates and stochastic fluctuations (supporting information).

Models of seasonal, viral respiratory illness demonstrate airborne respiratory virus transmissions exhibit a strong spatial decline (i.e. Gaussian) in the transmission rate with the distance between the recipient and the diseased host, and strongly depend upon local environmental factors such as air flow, temperature and humidity [14–18]. The strong dependence of the probability of transmission on host-recipient distance underscores the need to separate the data into pre-quarantine and post-quarantine periods, because the degree of social distancing changed dramatically post-quarantine. After quarantine, transmission rates decline in proportion to the strictness of the quarantine measures and the degree of compliance with them.

The transmission of a respiratory virus can be divided into four basic steps [19]: (1) a non-infected host interacts with the environment of an infected individual (2) the infected individual transmits *intact* virions to the non-infected host and (3) the virus infects the host (4) the virus replicates sufficiently for the host to become infectious.

In the first step, interaction, it is not necessary that the recipient and donor are in the infective environment simultaneously because, in a suitable environment, virus-droplets can remain suspended in the air for hours. Eventually, viruses inactivate due to the accumulation of environmental damage. This inactivation timescale governs the second step, which is viral transmission. The probability of infection, the third step, depends on the number of intact virions that deposit on the uninfected host and the susceptibility of the host. Susceptibility to infection depends on many factors, such as age, medical history, obesity and the internal humidity of the host, since a dry respiratory tract is more infection prone [20]. In the fourth step, replication, both demographic and environmental factors play a role. For example, temperature correlates with viral titer (hotter temperatures producing less titer) [15].

Consequently, the probabilities of these four steps are not independent. Environmental factors influence the frequency and conditions under which people associate, they regulate virus decomposition rates, modulate host susceptibility and severity of infection within the host. Droplet settling times, droplet evaporation times, and viral stability times, all depend on environmental factors such as humidity, temperature, wind etc.

Since all of these factors influence the transmission rate, they either must be included in the modeling, which introduces additional parameters and more complexity, or one can restrict the analysis to situations where the transmission occurs in environmental, social and economic steady-state conditions. The pre-quarantine Colombian data meets the steady-state conditions because of the lack of seasonality, the widespread daily use of transportation in densely packed buses and subways, the high population density in cities (table 1) and the suddenness and totality of the imposed quarantine.

## Materials and methods

### the drip model

A significant fraction of infective hosts of SARS-CoV-2 are asymptomatic [5, 6]. In addition, SARS-CoV-2 has a long incubation period of up to ∼15 days [21]. During this time, the disease host is infectious but asymptomatic. In Colombia, international travel was banned three days after the imposition of national quarantine [13]. Consequently, we approximate that the rate at which infectious travelers arrived in each city was roughly constant over the pre-quarantine period.

We model the daily arrival of SARS-CoV-2 into each city as a Poisson process with mean *I*. That is, we assume that each day prior to the quarantine, *I*[*t*] infected travelers arrive into a city where they begin infecting locals. We assume that on average, an infectious person infects *r* people each day, who in turn become infective (able to infect others) in one day. That is, on day *t* there are *I*[*t*] new infectious arrivals, as well as the *N* [*t*− 1] total infectious people from the day before, and the *rN* [*t*− 1] people they infected (Eq 1).

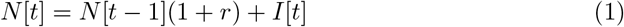

This difference equation is easily solved. We find that the expected number of infections on day *t* is:

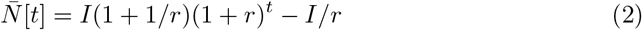

Eq 1 gives the expected number of infections in a given city on day *t*. In our analysis, we allow both *I*, the drip rate, and *r*, the transmission rate, to vary by city.

While the disease is spreading, an infrastructure is being established to detect the disease, which results in a dynamic disease detection rate. As a simple but useful case, consider a logistic increase in the detection rate. We define the total probability of detecting an arbitrary infectious disease host on day *t* as:

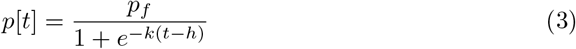

The detection rate increases from a small value (*p*(0) = *p*_*f*_ */*(1 + *e*^*kh*^)) at rate *k*, to a final detection capacity of *p*_*f*_ with half capacity reached on day *h*. The probability of detecting *c* cases of SARS-CoV-2 on day *t* is distributed as a Binomial distribution *B*(*N* [*t*], *p*(*t*), *c*(*t*)). The average (expected) number of infections detected on day *t* is then:

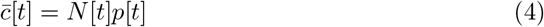

When *r* is small (which covers all cases of interest), the time-derivative of the log of the expected number of confirmed cases (the expected case log-velocity) is:

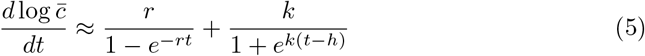

Note that the drip rate (*I*) and the overall detection rate (*p*_*f*_) have fallen out. Eq 5 is the basic equation that we use to fit the data. In the supporting information, we extend the drip model to include both varying drip rates and death and recovery.

### data

Covid-19 data for Colombia were downloaded from the web:

1. case data from the *Instituto Nacional de Salud* [22]
2. testing protocols from the *Ministerio de salud* [23]. Weather and population data were downloaded from:
3. World Weather Online [24]
4. Population data from City Population dot de [25]

Transportation data were taken from the web:

1. Airport data from the *Aeronáutica Civil de Colombia* [26]
2. Public transportation data from *Moovit* [27]

### data analysis

The complete data analysis procedure is as follows:

1. We began the analysis on March 10, 2020 four days after the confirmation of the first case in BogotáD.C.. We removed the first four days (which does not exclude any additional cases) to account for a large drop in testing that occurred during this time (Fig. 1).
2. In order to estimate the expected count number from the daily count number, we smoothed the daily confirmed case counts with a seven day triangle function. We include days past the quarantine cutoff in the smoothing to avoid edge effects from the filter. The data were then trimmed to the interval March 10 through April 3, 2020 for Bogotá D.C. and April 7 for the remainder of the cities.
3. We computed the count log-velocity (see supporting information) by first taking the numerical derivative of the smoothed count data (via the Python routine numpy.gradient) and then by dividing the gradient by the smoothed count data.
4. We fit the count log-velocity to the drip model according to the routine presented in the supporting information (Fig. S2).

Since the data span was about 30 days, exponential growth will only be observable for cities with transmission rates significantly greater than 1/30 *∼* 0.033. Consequently, 0.05 was considered to be the minimum threshold on the transmission rate to observe exponential growth in a given city. We associate transmission rates above this threshold with airborne transmission and transmission rates below this threshold with tourism and direct transmission [28].

In order to deduce the cutoff date for the quarantine, we examined the case counts in Bogotá (Fig. S1). Since we assume the transmission rate is constrained to be constant, the reduction in the transmission rate induced by the quarantine shows up in our model as a decrease in the detection rate (red vertical line). The vertical bar at April 3, 2020 denotes a combination of the impact of the quarantine on the spread of SARS-CoV-2 in Bogotá and a brief decline in the testing rate (Fig. 1). Since Bogotá went into quarantine 4 days before the rest of the country, its pre-quarantine data ends April 7, 2020.

## Results

When we apply our model to the data, the data only shows clear exponential growth in large cities with Relative Humidity (RH) over 80% and mean maximum daily temperatures below 30 degrees Celsius (Bogotá, Cali, and Medellín - Fig. 2). The RH in these cities is roughly the same (∼82 %), and they have similar total populations and urban population densities (table 1). The transmission rate declines among these cities with higher temperature.

**Fig 2.**
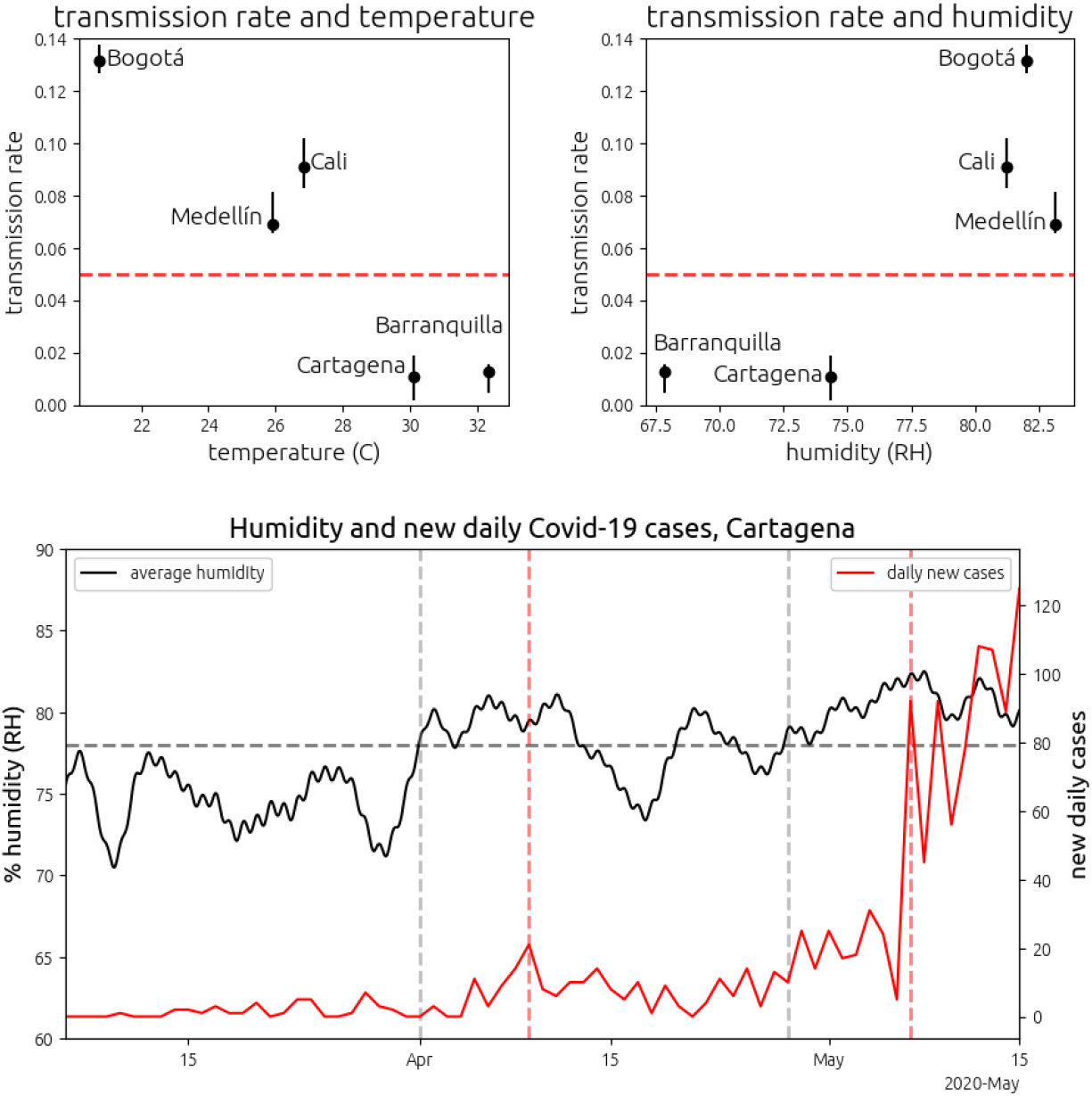
The transmission rate of SARS-CoV-2 with temperature and RH in the five largest cities of Colombia. (top) The dashed red line denotes a transmission rate of *r* = 0.05, which is our model’s threshold for observing clear exponential behavior and which we associate with airborne transmission. Note that these rates only apply to the pre-quarantine period. (bottom) Outbreak in Cartagena de Indias. The solid black line plots the average daily humidity in Cartagena (left axis). The solid red line plots the number of new cases of Covid-19 diagnosed in Cartagena (right axis). The horizontal dashed gray line plots a mean humidity level of 78%. The vertical dashed gray bar on the left denotes the first day with mean humidity at 78% since the first case of Covid-19 in Colombia. The first vertical dashed red line from the left, 8 days after the humidity increase, a spike in Covid-19 cases was registered. The second vertical dashed gray bar denotes the beginning of the second sustained period of humidity above 78% in Cartagena. A major outbreak began 9 days after the humidity rise.

We note that none of the smaller cities registered significant transmission rates in the pre-quarantine data at all temperatures and humidities. For example, Soacha and Bucaramanga compare to Bogotáand Medellín respectively. Soacha shares climate with Bogotá but has one tenth the total population (Soacha and Bogotá are neighboring cities and are connected by urban rail). Soacha has a higher urban population density than Bogotá. Bucaramanga has nearly one-fifth the population of Medellín (at similar temperature and humidity) and half the population density. Bogotá is more than 10 times larger than Soacha and Medellín more than twice as large as Bucaramanga. Moreover, Bogotá and Medellín are regional transportation hubs. Thus we conclude that the lack of transmission in Soacha and Bucaramanga is attributable to transportation factors as governed by total population and regional importance.

Although our analysis is focused upon pre-quarantine dynamics, there were two significant outbreaks post quarantine. The first was in Cartagena de Indias, a hot city on the Caribbean coast. While the weather in Colombia is nearly constant, there are minor temperature and humidity cycles associated with rainy and dry periods. The transmission rate in Cartagena correlates remarkably with sustained humidity above 78% (Fig. 2). We conclude from this that the outbreaks in Cartagena were driven by changes in RH, since temperature was nearly constant, and Cartagena had been under quarantine, isolated from international travel and under tight local travel restrictions for 50 days prior to the outbreak. A second outbreak in a small town in the Amazon Rainforest, Leticia, on the banks of the Amazon river, resulted in confirmed infections in roughly 1 in 20 residents, the highest confirmed per-capita infection rate in Colombia. This outbreak is remarkable because Leticia, as a small town, does not have an urban city center or large buses or subways, and the mean daily RH is around 94%, the highest RH for a city in this study.

## Discussion

Comodulation of viral infectivity by temperature and by relative humidity has been experimentally demonstrated in a variety of enveloped viruses [29] such as SARS-CoV [30], influenza [16, 31–34] and SARS-CoV-2 [35] (for temperature only) and other enveloped viruses [36] (Fig. 4). Three chief mechanisms have been proposed to explain the role of temperature and humidity upon enveloped viruses: *destabilization* of the virus within the droplet-matrix [31, 37], *evaporation and settling* of virus droplets [18, 37] and *reduced viral titer* produced by the host at higher temperatures [15].

**Fig 3.**
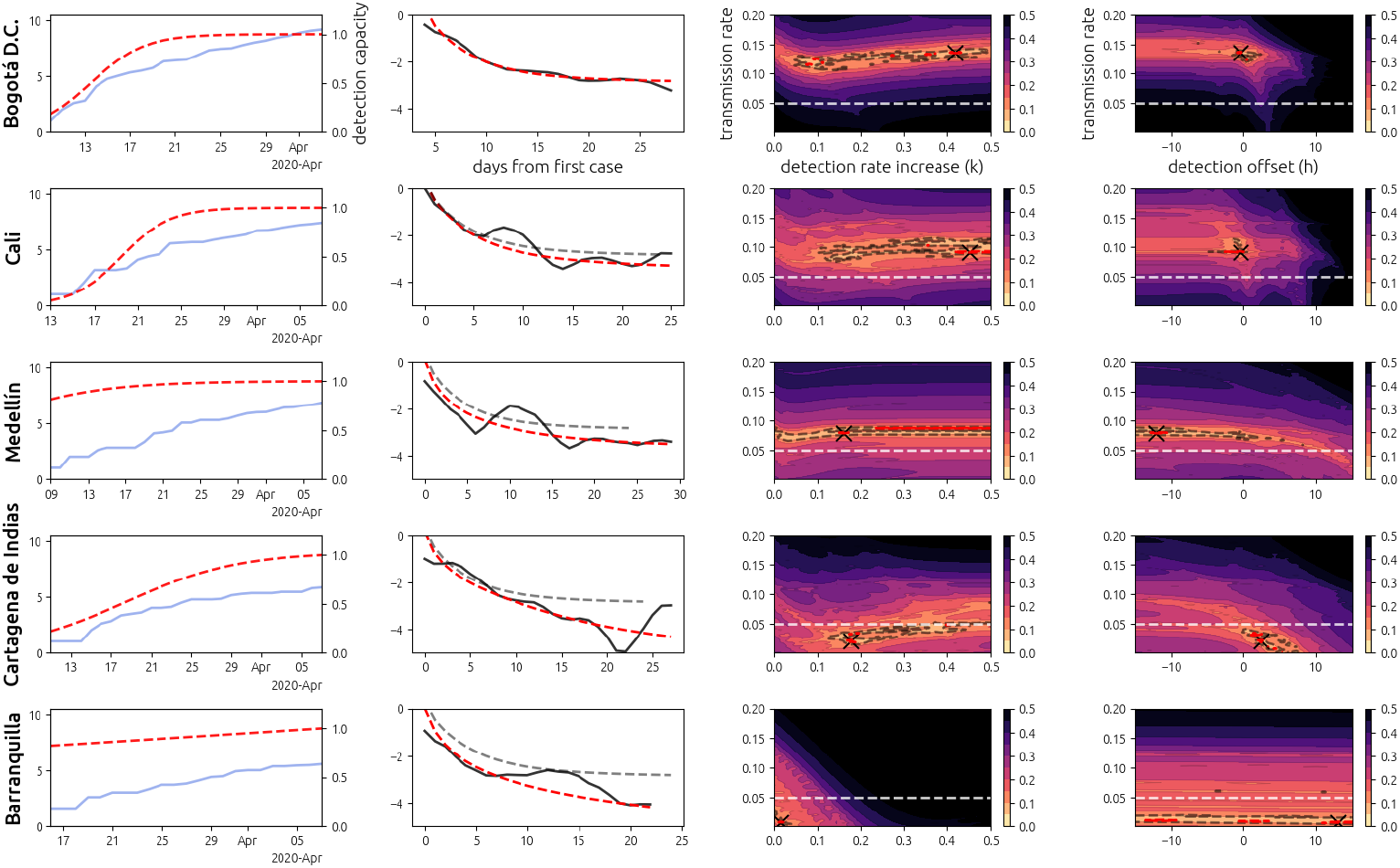
Covid-19 and model fit in the five largest cities of Colombia. Each row corresponds to a different city (left axis). The first column plots the cumulative number of confirmed cases of Covid-19 over the pre-quarantine period (blue) as well as the inferred detection capacity within the city (dashed red, right axis). The second column plots the log-velocity (see supplementary material) of the data (black) and of the model fit (dashed red), with the model for BogotáD.C. shown for reference (dashed gray). The third and fourth columns show cross-sections of the error function *e*(*r, k, h*) between the model under parameters (*r, k, h*) (transmission rate, detection rate increase rate, and detection rate half capacity date). The error bars are given in terms of percent increase from the global minimum. Global minima are denoted by a black cross. The dashed white line indicates the threshold *r* = 0.05 for observing clear exponential growth in our model. The red contours denote an increase from the global minimum of 5% and the gray contours denote an increase from the global minimum by 10%.

**Fig 4.**
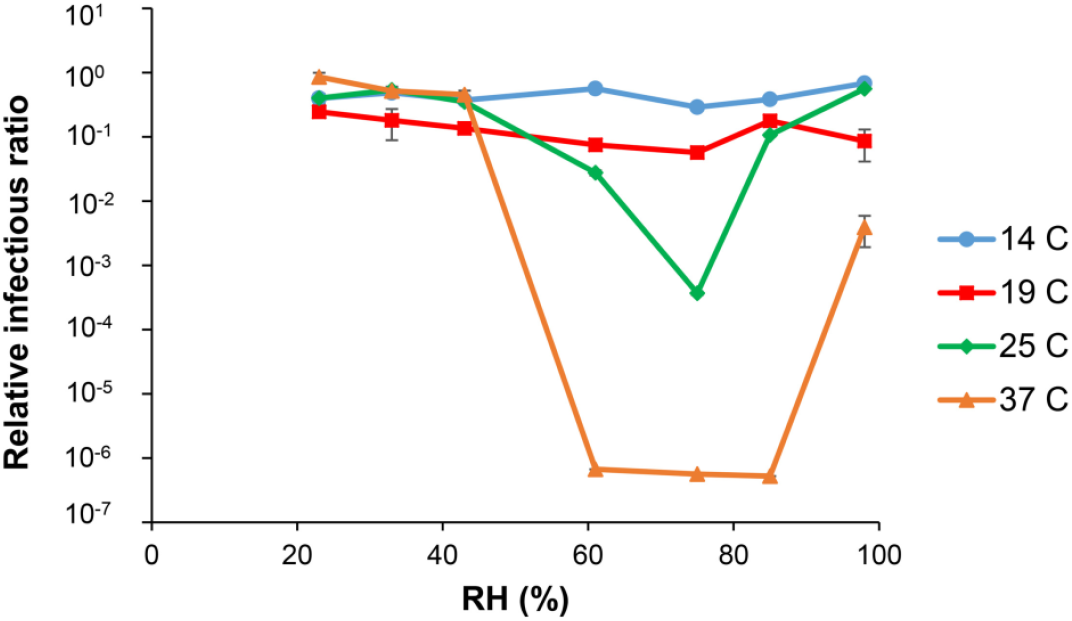
Phi6 infectivity with temperature and RH. Original data from Prussin et al [36] on the relationship between temperature, RH and Phi6 infectivity. Phi6 is an enveloped virus used as a model for influenza, coronavirus and other respiratory viruses. The error bars are shown but too small to be visible in the figure.

Reduced viral titer could account for some of the reduced transmission observed at higher temperatures. For influenza, Halloran et al. [15] show a reduction in peak nasal titer for influenza by an order of magnitude for temperatures between 5 and 30 degrees Celsius in guinea pigs [15]. However, viral titer cannot explain the outbreak in Cartagena which appears to be driven entirely by high RH, since the air within the human respiratory system is saturated and unlikely to vary with small changes in ambient RH [38].

Although direct measurements at short timescales in conditions that mimic respiratory droplets are unavailable [37], temperature driven destabilization in solution apparently occurs on the timescale of minutes [31, 35, 36]. Droplet settling timescales vary from minutes to hours depending on droplet size [37]. Given the dense packing on public transportation in cities with attenuated transmission, we reason that the mechanism behind this attenuation must act on the timescale of seconds [15, 39] to be responsible for low infectivity. This rules out droplet settling and temperature driven viral destabilization as the cause of lower infection rates in these areas.

Airborne respiratory droplets evaporate down to half their size in about one second [32, 37]. This evaporation is thought to influence viral stability through salt and protein concentrations, pH gradients, and surface sheering [32, 37]. RH and temperature both influence droplet evaporation, with temperature influencing the rate and RH determining the final droplet size [15]. While the timescale of droplet shrinkage has been studied, the timescale of viral inactivation within the shrunken and toxic respiratory droplets is, to our knowledge, unknown [37]. Determining this timescale could have important implications for policy surrounding the Covid-19 pandemic, since fast destabilization at specific temperature and humidity intervals would have both prevention and therapeutic implications. Additional experiments are necessary to resolve the impact of temperature and humidity on the infectivity of SARS-CoV-2 within respiratory droplets.

## Conclusion

Our observations suggest a decline in the spread of SARS-CoV-2 with temperature in regions with moderate humidity. We predict an increased probability of outbreaks as the relative humidity approaches and surpasses 80% even at high temperatures. Cities near large bodies of water, such as Beijing, where the humidity rises to around 80% in the summer, are at particular risk for outbreaks as the Prussin data suggests an exponential rise in infectivity with humidity at warm temperatures (Fig. 4). Given the crowded public transportation systems and high urban population densities of cities with highly attenuated transmission, we hypothesize that the attenuation is caused by rapid viral inactivation within the respiratory droplet matrix as mediated by evaporation through temperature and RH, with direct temperature effects upon the virus contributing. Experimental confirmation of this hypothesis could have significant implications for policy surrounding the Covid-19 pandemic. For example, indoor climate control (or lack thereof) might be considered as a means of mitigating the spread of SARS-CoV-2. The same mechanisms of viral destabilization within evaporated respiratory droplets could be considered as a means of directly combating the virus. Establishing the timescale of viral destabilization within respiratory droplets, resolved on the shortest timescale possible, may then provide important information about the biology and transmission mechanisms of SARS-CoV-2.

## Supporting information

As described in the main text, it is essential to distinguish the *transmission rate*, or the propensity of the disease to spread, from both the total disease prevalence and the number of confirmed cases in a given population. Here we clarify the notions of detection rate, testing rate, and drip rate and quantify their impact upon the scaling of the confirmed case number and the apparent transmission rate.

The *detection rate* describes the total probability of detecting a case of SARS-CoV-2 in a given population. This rate undoubtedly increased over the period covered by our analysis since the testing rate increased by over two orders of magnitude during that time (Fig. 1). In section *detection rate scaling*, we show that the detection rate introduces an additive factor into the exponent of the expected number of confirmed cases.

In section *drip rate scaling*, we show that the *drip rate*, or the rate at which infected travelers arrive into a city, introduces a multiplicative constant into the total number of infectious hosts in a given city which translates to a multiplicative constant in the expected number of confirmed cases.

In section *testing is driven by perception*, we demonstrate that the testing rate is not equivalent to the *detection rate*, since the testing rate is driven by human decisions about going to the hospital, ordering tests, and testing protocols, whereas the *detection rate* describes the final probability of detecting a case of Covid-19. As an example, we show that panic can drive a wave of non-infected people to receive tests, driving up the testing rate, without changing the overall detection rate (subsection *panic in a disease free population*).

We make a distinction between the testing rate and the detection rate, because we use the observed exponential growth in the testing rate as motivation for including the detection rate in our data model, but do not introduce the testing data into our data analysis as a constraint, because our algorithm deduces the detection rate from the data without the need for constraints. Furthermore, our analysis shows that we should not expect the testing rate and the detection rate to have more than qualitative agreement.

In section *count log-velocity*, we show that the log-velocity, or the time-derivative of the log of the expected number of confirmed cases, is a useful measure of disease spread because it is independent of the drip rate and the overall detection rate. Since the log-velocity still depends on the detection rate dynamics, we include a logistic model for detection (section *drip model*).

### detection rate scaling

Consider a population with a constant number *N* cases of Covid-19 with the total probability of detecting a case of Covid-19 written as *p*[*t*]. Since the probability of detecting *c* cases of Covid-19 is a binomial in the *N* cases, the expected number of *confirmed* cases on day *t* is:

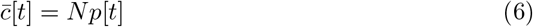

When the detection probability is small, *p* ≪ 1, the fluctuations in the observed case number are of order:

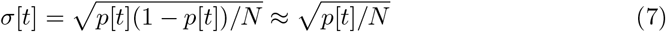

Note that both the expected number of cases, 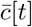, and the fluctuation in the number of cases, *σ*[*t*], are *increasing functions of the detection rate p*[*t*]. That is, both the measurement mean and the measurement variance depend on the detection rate. We assume that the function *p*[*t*] can be well approximated by a logistic function. Hence,

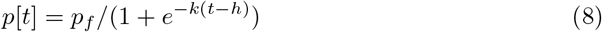

Here the detection rate converges to a final rate *p*_*f*_ with rate *k* and reaches half the final rate on day *h*. In the early stage of the pandemic, while the detection capacity is a small fraction of its later capacity, we have *t < h* and exp{*k*− (*t* −*h*)}≫ 1. Then, to first order:

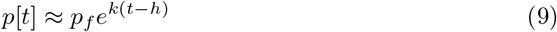

The log of the expected number of cases on day *t* is then:

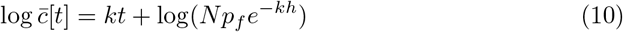

Eq 10 shows that the log of expected number of counts increases linearly with rate *k*. Thus, at the beginning of testing ramp-up, *constant infections with logistic growth in the detection rate is indistinguishable from exponential growth in the number of infections with a constant detection rate*. To see this, simply substitute *N* = *Ne*^*rt*^ for the number of infections and substitute *p*(*t*) = *p*_*f*_ for the detection rate. Then

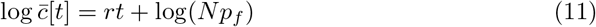

Since *p*_*f*_ is initially unknown, the observed case dynamics will appear equivalent when the number of infections is constant and the detection rate grows logistically versus when the number of infections grows exponentially and the detection rate is constant.

### drip rate scaling

In the previous section we illustrated that the local detection rate introduces an additive factor into the apparent transmission rate of the disease. In this subsection, we demonstrate that the drip rate introduces a multiplicative factor into the total number of infections in a given location under very general assumptions. *We only assume that in the early stage of the disease, the disease hosts are weakly-interacting*.

Under the weakly-interacting disease host assumption, the infected disease travelers who arrive in a given city create new pockets of infection. Since we assume that the disease hosts are weakly-interacting, a city with twice the drip rate will have twice the number of pockets in which the disease grows. As long as the weakly-interacting assumption is true, these pockets grow independently at the same rate, with no overlap. The total number of infections will then be the sum of infections over each of these disease pockets. In this way, the drip rate introduces a multiplicative factor into the expected number of infections regardless of the details of how the disease spreads.

We quantify this intuition in the following way. Let each person in a given location (i.e. city) be assigned a unique number 1, 2, … *N* for *N* total people. Let *T* [*t*] be the set of diseased travelers (*τ*) that arrive in this location on day *t* with *I*[*t*] = |*T* [*t*]|. Then *T* [1] = {*τ*_11_, *τ*_12_, …, *τ*_1*I*[1]_ } and *T* [2] = {*τ*_21_, *τ*_22_, …, *t*_2*I*[2]_ }. On average, each of the *I*[*t*] travelers that arrive on day *t* infect *r* ≪ 1 people per day. That is, on average every 1*/r* days, each traveler will infect one new person, and every subsequent 1*/r* days this new infectee will go on to infect another person, and so on. Consider the function *λ*_*τjt′*_ [*t*] that returns the unique identifier(s) of the person(s) infected by traveler 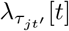 on day *t* (i.e. the *j*-th traveler to arrive on day *t*). Note we are only counting people who live in the location to be infectees and are not counting other travelers that the traveler infects. Then the function 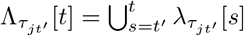 denotes all people that the infected traveler *τ*_*jt*_*′* has directly infected until day *t*.

Since we are considering the beginning stages of the disease and are neglecting re-infection, 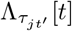 must be disjoint for each of the travelers. That is, without reinfection, a person can only be infected by one traveler. Next, we consider the pool of people with whom each infectee interacts. Denote by *c*_0_[*n*] all the people whom person *n*, not a traveler, can potentially infect by direct interaction. These are people who are susceptible to the disease and who come into contact with the infectee (*n*). Now denote by *c*_1_[*n*] all of the people whom person *n* can potentially directly infect. That is, *c*_1_[*n*] is the union of *c*_0_[*n*] with each *c*_0_[*n*] for each of the people *n* in *c*_0_[*n*]. In this way, we can consider *c*_2_[*n*], *c*_3_[*n*] and so on, each of these sets becoming larger and larger as the pool of potential infectees becomes larger and larger. We refer to each stage of disease transmission as a generation. That is, the people in the set 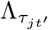, each infected by traveler *τjt*′ between day *t* and *t*, are the first generation of infectees. Then the people that these infectees infect are the second generation, and so on.

For the early stages of the disease, we need not consider very many generations. The generation timescale is 1*/r* and so the time for the disease to progress to the k-th generation is 1*/r*^*k*^. For *r* ≪ 1, this time quickly becomes very large.

Thus, our assumption of weakly interacting hosts amounts to the assumption that the number of generations is small and the social circles of the early generations of infectees from each traveler are approximately disjoint. Recall that the first generation of people infected by traveler *τjt*′ until day *t* is 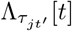. Then the total set of people who are susceptible to a second generation infection is 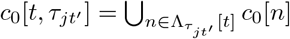 and the total set of people who are susceptible to a third generation infection from traveler 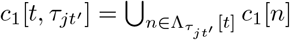 and so on.

Eventually, |*c*_*m*_[*n*]| = *N* and the extended social circle of an arbitrary person includes the entire city. But for *m* small, we assume that the social circles are approximately disjoint, meaning that the presence of more infectious travelers does not restrict the dynamics of the generations of infections that stem from each traveler. Let *m* be the maximum number of generations to which the disease spreads during the early stages. We assume that |*c*_*m*_[*t, τjt*′] ⋂*c*_*m*_[*t, τ*_*kt*_*II*]| ≈ 0: the set of potential infectees descendant from each traveler, over the relevant number of generations, is approximately non-overlapping.

Under this assumption, the presence of more infectious travelers does not restrict the spread of the disease. Thus, at any given time, a city with twice the drip rate will have roughly twice the total number of infections as a city with the same transmission rate and half the drip rate. This is what we mean by the statement that the drip rate introduces a multiplicative constant into the total number of infections. We can express this symbolically as follows. Denote by *N*_0_[*t*] the average disease dynamics subsequent to the infection of one local resident (*N*_0_[0] = 1) and denote by 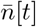 the number of infections that the average traveler spreads per day. Then the total number of infections on day *t* is roughly:

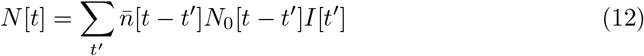

The expected value of this this quantity is easily seen to be:

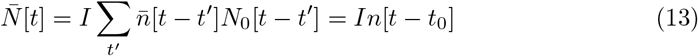

Here we see that a multiplicative constant has been introduced. Note that we have not introduced a specific model for how the disease spreads. We have only assumed that the disease spreads independently.

From this we conclude that the total number of cases in a given city does not necessarily inform us about the propensity of the disease to spread in the environment of the city, especially for short timescales and in cities where the transmission rate is low. As we will show in the next section, *the dynamics of the count numbers* tell us more about the transmission rate than the absolute scale of the number of infections at any time.

### count log-velocity

So far we have only considered (*k*), the rate at which the disease detection probability increases, and the drip rate (*I*), the rate at which the disease is introduced into a city. We have not yet considered the total probability of detecting a given case of Covid-19 at a given moment in time.

Consider two cities, city 1 and city 2, that are alike in all respects except for the overall detection rate *p*_*f*_. Also assume that the disease transmits in both cities at the same rate (*r* = *r*_1_ = *r*_2_), the disease is introduced into the two cities at the same rate (*I* = *I*_1_ = *I*_2_), and the disease detection infrastructure grows at the same rate (*k* = *k*_1_ = *k*_2_).

It is not necessary to specify the model of the disease dynamics, as long as the assumption that the disease spreads independently from each infected person is true. There may be other parameters of the model that we do not specify here. Whatever these may be, they are assumed to be equal for the two cities. Likewise, we do not specify a model of the detection rate increase in the two cities, we only assume that it is some function *f* (*t, k*) such that

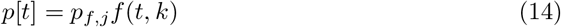

There are no restrictions on *f* other than it depend upon some rate *k* parameter that governs the rate of change of *p*[*t*]. The function *f* (*t, k*) could depend upon other parameters which we have not specified but which are assumed equal between the two cities.

The probability of detecting *c* cases of Covid-19 in either city is a binomial and thus the expected number of counts is (from Eq 15):

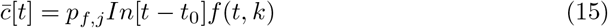

Eq 15 explains why static count numbers convey little information about the transmission rate. Depending upon the total detection probability *p*_*f*_ and the drip rate, *I*, the total count number between two otherwise identical cities could vary (hypothetically) by an order of magnitude or more. Note that both *p*_*f,j*_ and *I* fall out of the time derivative of the log of the expected number of cases :

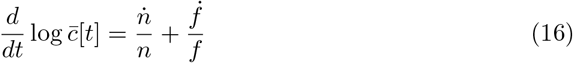

### testing is driven by perception

In Colombia, From February to April 2020, detection was conducted at hospitals on symptomatic patients. In order for a patient to be diagnosed as Covid-positive, three steps were required: (1) a symptomatic disease host must go to the hospital, (2) the attending physician must order a test and (3) the test must result positive.

These three steps are not independent. For example, a severely symptomatic host is more likely to go to the hospital and request attention than a weakly symptomatic host or an asymptomatic host. Likewise, a severely symptomatic host is more likely to have a doctor order a test and more likely to test positive for Covid-19 than a moderately symptomatic patient.

Consider the following simple model for testing. The expected number of *unique* tests conducted on day *t, N*_*T*_ [*t*], as the product of the number of people in the city times the probability of being tested. Since being tested requires going to the hospital, we separate the probabilities by the product rule:

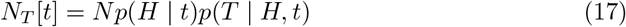

This division is essential because it divides the dynamics into two distinct populations: the general population and the hospital population. The general population dynamics *p*(*H*| *t*) are driven by self-perception of symptoms, whereas the hospital dynamics *p*(*T*| *H, t*) are driven by physician perception of symptoms and hospital protocol.

Note that the number of tests conducted on a given day is not, in general, equal to the number of *unique* tests conducted on a given day. This is because most countries implement protocols that call for duplicate testing. For now, we will not consider duplicate testing since we are only interested in the basic formulation and scaling of the testing dynamics.

Here *p*(*H*| *t*) denotes the probability of going to the hospital with Covid-related symptoms, which for now, are assumed to be the only requirements for a symptomatic person to be tested. We expand the probability of going to the hospital in terms of those who are infected (*i*) and those who are not infected (*ī*) with SARS-CoV-2: those who are not infected include those who have other infections, e.g. influenza.

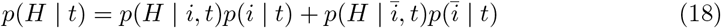

Note that many people who are not infected with SARS-CoV-2 will have symptoms consistent with Covid-19. These symptoms may result from (1) other respiratory viruses (2) other health problems and (3) mass psychogenic illness. It is well documented in psychology literature that the suggestion of symptoms provokes symptoms in a large percentage of the population, particularly when group consensus is involved [40].

We further expand each of the hospital terms in Eq 18 in terms of the severity of the symptoms. We use a three-grade ranking of symptoms: asymptomatic (0), mildly symptomatic (1) and severely symptomatic (2) (see table S1). We assume that only symptomatic patients show up at the hospital as Covid-19 testing candidates, and that the general population dynamics is driven by self-assessed symptoms (such as difficulty breathing and fatigue).

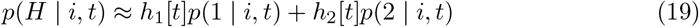

The term *h*_1_[*t*] = *p*(*H*| 1, *t*) (*h*_2_[*t*] = *p*(*H* |2, *t*)) denotes the probability of going to the hospital given grade one or grade two symptoms. Here *p*(*H* |1, *i, t*) = *p*(*H* |1, *t*). This is because the probability of going to the hospital cannot depend on the true disease status as the decision to go to the hospital is subjective: the individual is not certain of his/her actual disease status.

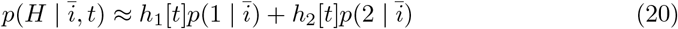

Combining and re-arranging terms:

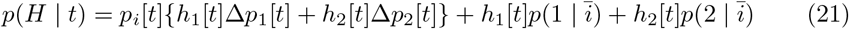

HereΔ*p*_1_[*t*] = *p*(1| *i*) − *p*(1| *ī*) is the difference in the probability of exhibiting grade one symptoms in the infected population (*i*) and the remainder of the general population (*ī*);Δ*p*_2_[*t*] has an equivalent definition for grade two symptoms and *p*_*i*_[*t*] denotes the disease prevalence among the population.

Eq 21 shows that the general population dynamics is driven by two terms: an infected sub-population term and an uninfected sub-population term. Interestingly, the infected sub-population term is driven by the fraction of people infected in the total population, *p*_*i*_[*t*], and *the difference in symptom rate* between those who are infected *I* and those who are not infected *ī*. That is, if the rate of moderate and severe symptoms between those who are infected and those who are not infected were the same, the number of people who show up at the hospital would not change as the disease spread.

Whereas patient perception drives general population dynamics, physician perception drives hospital dynamics. We now expand the probability of being tested in terms of the severity of symptoms under the assumption that the physician does not administer tests to asymptomatic patients and always administers tests to severely symptomatic patients:

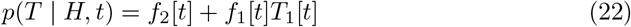

with *f*_2_[*t*] = *p*(2 |*H, t*), *f*_1_[*t*] = *p*(1 |*H, t*) and *T*_1_[*t*] = *p*(*T*| 1, *H, t*). In other words, *f*_2_ is the fraction of people at the hospital that the physician diagnoses as having grade two symptoms, *f*_1_ is the fraction of people at the hospital that the physician diagnoses as having grade one symptoms and *T*_1_[*t*] is the probability of administering a test to a grade one symptom patient (as diagnosed by the physician).

Note that we are suppressing an important condition here for the sake of compact notation. Here *p*(2 | *H, t*), for example, denotes the fraction of people at the hospital with grade two symptoms *according to the physician’s perception*. We might denote this with a *P* for emphasis - e.g. *p*(2 | *H, t*) = *p*(2 | *H, t, P*). We emphasize this distinction because it is tempting to apply Bayes’ theorem to simplify these expressions, but doing so requires care and keeping track of the conditions that we have largely suppressed in this brief presentation.

We note two interesting cases.

### panic in a disease free population

When there is no disease within the population, then *p*_*i*_[*t*] = 0 for all times considered. In that case, the probability of going to the hospital among the general population is (from Eq 21):

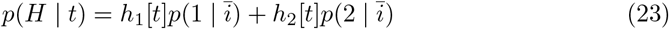

A rough upper bound order of magnitude estimate for the probabilities *p*(1| *ī, t*) and *p*(2| *ī, t*) is about 1% and 0.1% respectively. Consider that during a typical flu season, over 50 million Americans contract the flu. For a population of 350 million, that is one in seven. Given that peak flu season spans about three months, then order 0.1% of the population is infected per day (new infections) during the flu season. If we make the approximation that all infections are symptomatic and the symptoms last for five days, then about 0.5% of the population will have symptoms *only from influenza* consistent with Covid-19 on a given day of peak flu season. Further note that roughly 10% of the population goes to the hospital per year, or about 0.02% per day assuming a uniform distribution. Thus, if we have *h*_1_∼ 0.01 and *h*_2_ ∼0.1 we obtain the right order of magnitude for people going to the hospital.

The small order of these numbers is important because a jump in *h*_1_[*t*] or *h*_2_[*t*] or in the probability of perceiving symptoms (*p*(1 | *ī, t*) and *p*(2 | *ī, t*)) can provoke a hospital rush. Recall that *h*_1_[*t*] is the probability of going to the hospital with moderate symptoms. As a crude but simplifying assumption, we make the approximation that in a panic, all people with moderate or severe symptoms will go to the hospital: *h*_1_ and *h*_2_ will both be of *O*(1).

This, in the panic caused by the introduction of a new disease, the fraction of people in the general population who experience moderate symptoms will make a large jump. In numerous studies in the literature, mass psychogenic illness (MPI) has been shown to arise and give rise to real, measurable symptoms, even when there is no underlying disease. This phenomenon is more prevalent in women than men (particularly young women). There were several instances of young women spreading panic on social media in the USA when they did not have the disease (see, for example, [41]).

As a model of MPI, we introduce the following form for the spread of psychogenic symptoms:

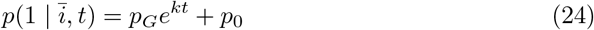

Here *p*_*G*_ ∼1/*N* ≪ *p*_0_ since the psychogenic symptoms usually begin with a single case. This model is consistent with numerous case studies that show a rapid spread of symptoms among peer-connected groups. In some cases, an entire factory or an entire military base is crippled overnight by the spread of spurious symptoms. Indeed, the only factor slowing down the spread of psychogenic symptoms is (1) the delay in the transmission of information among susceptibles and (2) the number of people in social groups, as proximity of a symptomatic appears to be a key factor in triggering psychogenic symptoms.

For simplicity, we are limiting psychogenic symptoms to moderate symptoms. We assume that for all times considered, including *t* = 0, *p*(1| *ī, t*) ≫ *p*(2| *ī, t*), or that cases with moderate symptoms greatly outnumber cases with severe symptoms. Then to first order, the probability of going to the hospital is:

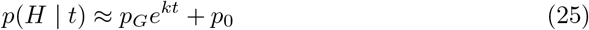

The total number of unique tests on day *t* is then:

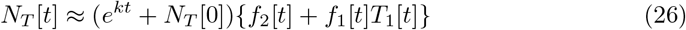

Recall that *T*_1_[*t*] is the probability of a test being administered to a person with moderate symptoms. We assume that under a panic, *T*_1_[*t*] →1. In such a situation, the testing rate becomes

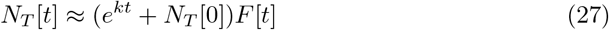

where *F* [*t*] is the total fraction of the hospital patients that the attending physicians perceive as having symptoms (moderate or severe) consistent with Covid-19. In the early stages of a panic, *F* [*t*] must be rapidly increasing, or perhaps constant if the medical staff is particularly stoic. In any event, *F* [*t*] is non-decreasing. Thus, MPI can cause an exponential rise in the testing rate, even when there is no disease in the population. In such a situation, for any significant false positive rate, the number of positive test results will also exhibit exponential growth.

### ignorance among a diseased population

Now consider the opposite case: a disease spreads rapidly throughout a population who fail to recognize the outbreak. Assume exponential growth of the disease prevalence in the population *p*_*i*_[*t*] = *p*_0_*e*^*rt*^ and assume that people with moderate symptoms do not go to the hospital *h*_1_[*t*] = 0. Since there is no awareness of disease spread, *h*_2_[*t*] = *h*_2_ is a constant. Then the probability of going to the hospital is:

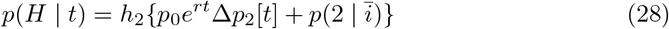

We assume that *p*_0_ ≪ *p*(2| *ī*) so that at *t* = 0 the vast majority of people with severe symptoms do not have the disease. Then we can consider two times, *t* ≪ *T* and *t* ≫ *T* with *T* = *r*^−1^ ln(*p*(2 | *ī*)*/p*_0_). In the first case, *t* ≪ *T*, the fraction of people going to the hospital is constant at ∼ *h*_2_*p*(2 | *ī*). Then for times *t* ≫ *T*, the fraction of people going to the hospital grows exponentially ∼ *h*_2_*p*_0_Δ*p*_2_[*t*]*e*^*r*(*t*−*T*)^.

Since there is no change in disease awareness, the fraction of the people showing up at the hospital getting tested is a constant. That is *p*(*T* |*H, t*) = *F*_0_. Then for times *t* ≪ *T* the fraction of the population tested is:

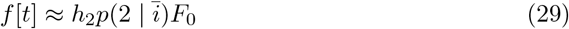

We have now shown that, at least for some period of time, the testing rate can be constant in a region of exponential disease growth and that, conversely, the testing rate (and potentially the number of disease diagnoses) can grow exponentially in a region where there is no disease presence. On this basis we assert that disease testing rates are driven by perception. Likewise note that the disease testing rate cannot be considered proportional to the disease prevalence rate *p*_*i*_[*t*].

The model in this section is simple. The main purpose of this section is to show that testing represents a complex and dynamic social phenomenon and (2) testing cannot be considered to be proportional to disease prevalence and (3) testing is not proportional to the probability of detecting a disease host in a given population.

### continuing the drip model

In (S4 fig), we plot simulations for the drip model presented above in Materials and methods. We estimate the standard deviation in the count number as:

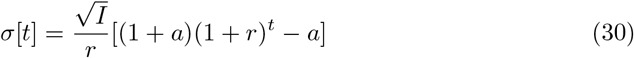

Here *a*∼ 0.15 is a numerical parameter. Eq 30 shows that the fluctuations in the count number vary exponentially with time. This is another reason to distrust absolute count numbers without dynamics: stochastic fluctuations alone can produce exponential variations in count number.

### varying drip rate

The dynamics can be expressed purely in terms of the drip rate as follows:

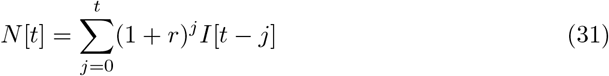

From Eq 31 we see that (1) the early drip rate drives the dynamics since these have the largest weighting and (2) the number of infections is linear in the drip rate.

### recovery and death

We now extend the drip model presented above to include recovery and death. We refer to this as “removal” of the infectious host or “resolution” of the infection. As of the time of writing, the distribution of recovery times for SARS-CoV-2 is not precisely known, but it varies between one week and one month and depends upon various factors such as host immune response. We model removal by subtracting a removal fraction from the number of infections on day *t*:

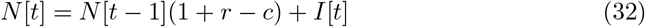

Here *c* is the fraction of people in the pool of infectious hosts who recovered or died. This fraction *c* depends upon (1) time (2) the transmission rate *r* and (3) the distribution of removal times. We will clarify each of these points in what follows.

According to the drip model, the number of new infections added to the population on day *t* is:

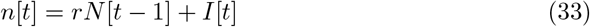

The number of people that are removed from the pool of infectious hosts (i.e. the number of people who recover or die) on day *t* follows some unknown distribution of removal probabilities. Let *q*[*t*] be the number of people removed on day *t*. Then:

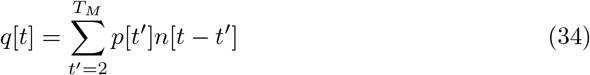

Here *p*[0] = *p*[1] = 0 since we only count infections that are infectious for at least one full day and *T*_*M*_ is the maximum infection duration. Note also that we are assuming that the travelers are recently infected. From Eq 32, we see that the fraction *c* on day *t* is defined as the ratio of the number of people removed on day *t* to the total number of infections on day *t* − 1. That is

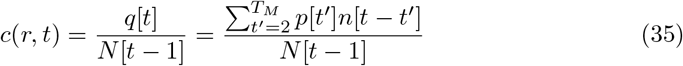

From Eq 35 we observe the dependence of *c* upon the transmission rate *r*, the time *t*, and the distribution of removal times *p*[*t*].

We estimate the initial order of *c* as follows. First, we approximate the distribution *p* in Eq 34 by a delta function. That is, we suppose that *p*[*T*] = 1 (i.e. all infections are resolved on day *T*). Then on day *t* = *T*, the first removals leave the infectious pool. The fraction of people that leave can be approximated as the ratio of the expected number of removals on day *t* = *T* to the expected number of infectious hosts on day *N* [*T* − 1]:

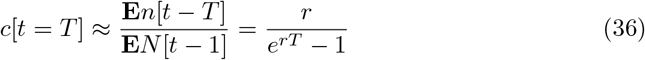

This equation is only valid at *t* = *T*. If *c* ≪ *r*, then this estimate will remain valid for future times. Using estimates on the transmission characteristics of SARS-CoV-2 by one of our coauthors, we obtain an estimate for *T* = 15.5 days [42]. With this long infectious period, the impact of including recovery dynamics on the early spread within Colombia is not significant (S5 Fig.).

## Data Availability

All data is publicly available.

https://www.ins.gov.co/Paginas/Boletines-casos-COVID-19-Colombia.aspx

https://www.worldweatheronline.com/

https://citypopulation.de/

## Acknowledgments

GB’s research is partly supported by ORIEN/NOVA, the Department of Defense and NIH/NCI. GB thanks the University of California at San Diego for hospitality during his sabbatical year 2019-2020.

---

**Fig S1.**
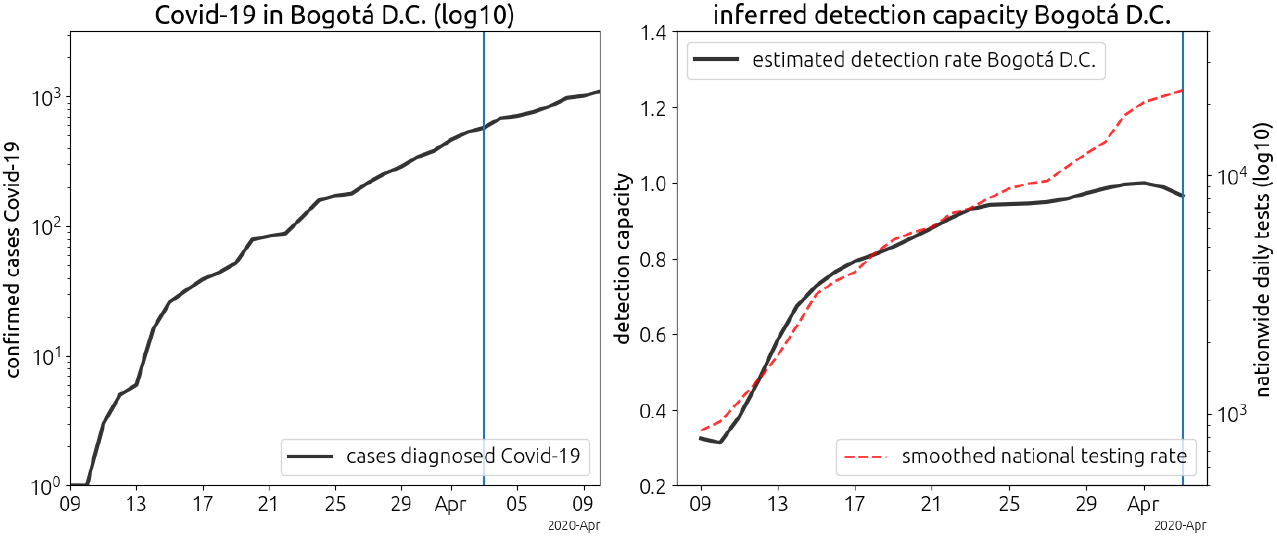
Covid-19 and testing in Bogotá D.C. In the right panel, the national testing rate (dashed red, log base 10) and inferred detection capacity for Bogotá D.C. only (black). Note we have plotted the smoothed national testing rate in dashed red; this plot is on a log10 scale and only serves to show qualitative agreement between the inferred detection rate and the daily testing rate. The national testing rate includes testing for all cities in Colombia.

**Fig S3.**
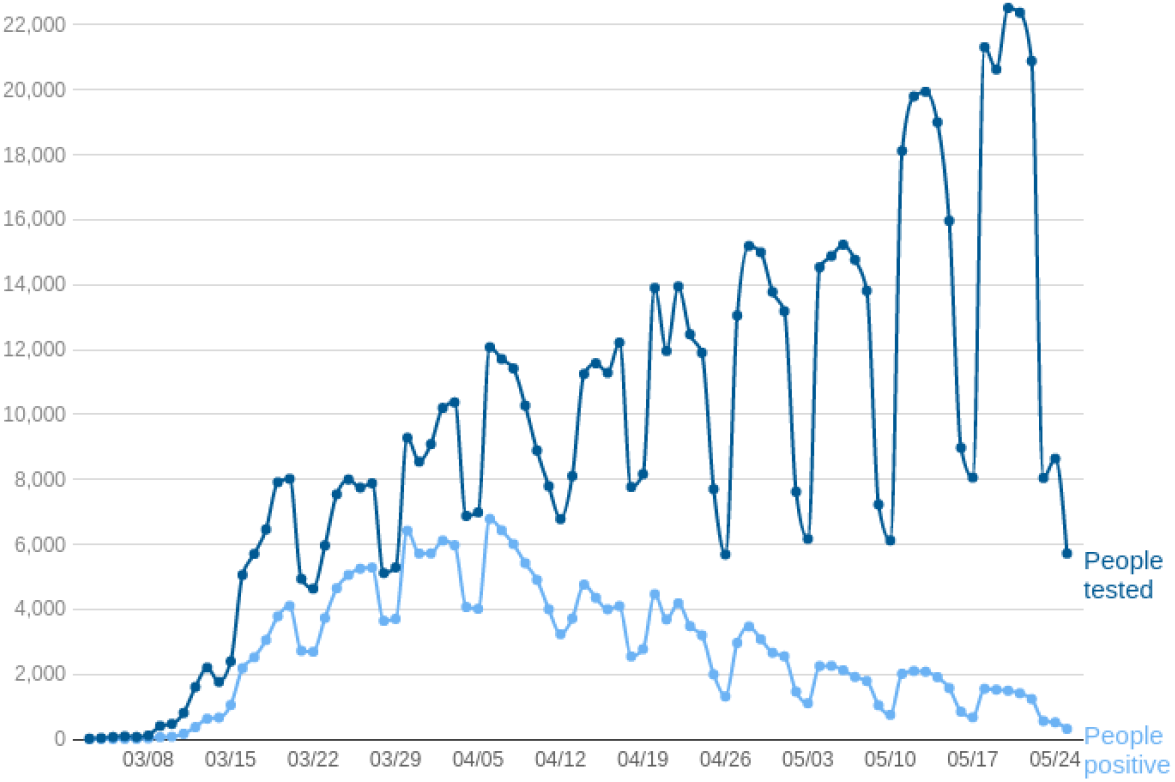
Daily testing data from New York. Figure from the official website of the city of New York [43].

**Fig S4.**
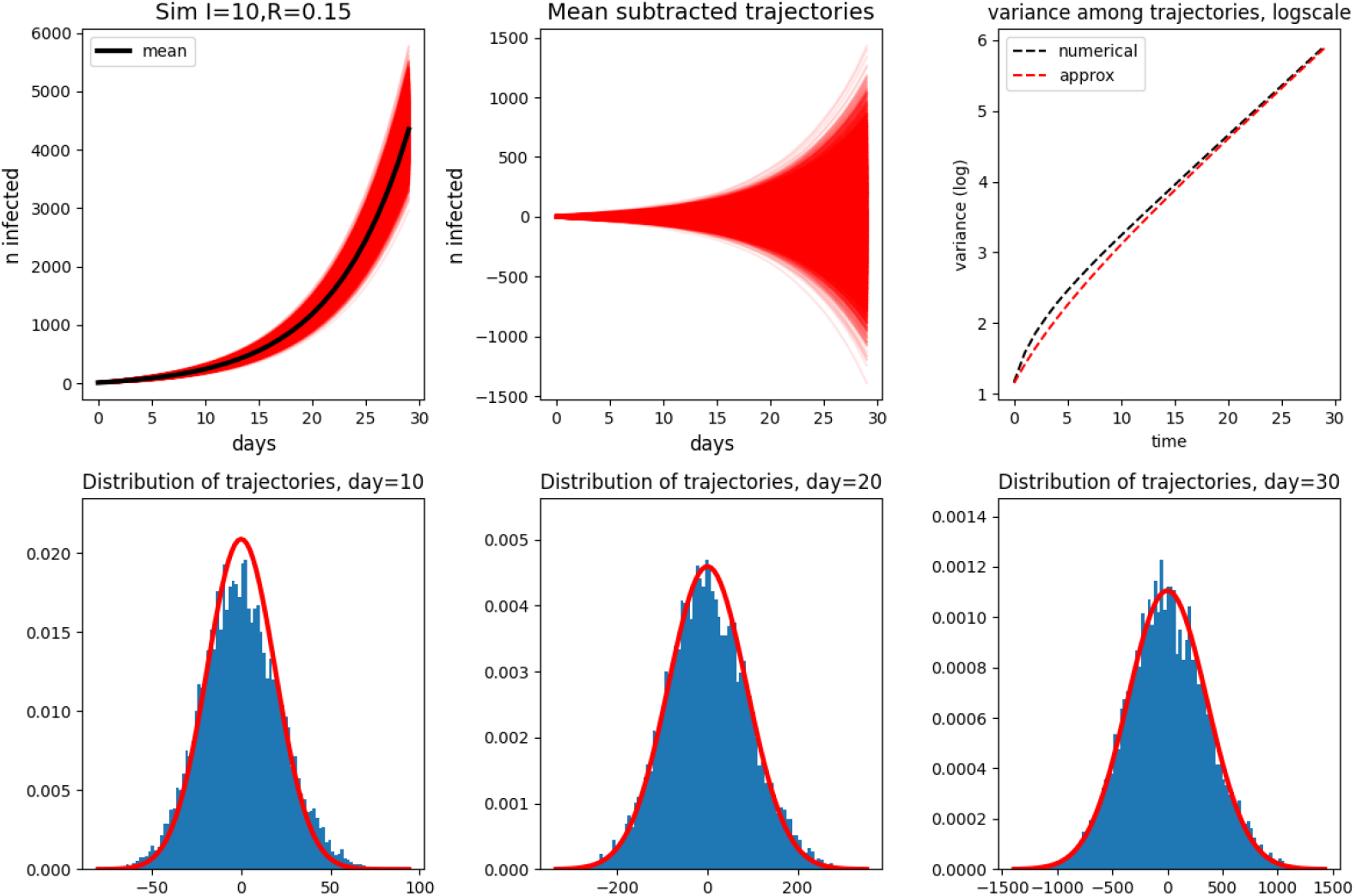
Drip model simulation. Here *I* = 10 and *r* = 0.15. In the top row we plot (left panel) the trajectories (red) with the mean trajectory in black (computed from equation 2, (center panel) the mean-subtracted trajectories, (right panel) the standard deviation as computed numerically (black dashed line) and by equation 30 (red dashed line). In the bottom row, with plot the distribution of trajectories at day 10 (left), 20 (center) and 30 (right). The Gaussian fit in bold red in each of the plots in the bottom row is obtained by applying the mean from equation 2 and the standard deviation from Eq 30.

**Fig S5.**
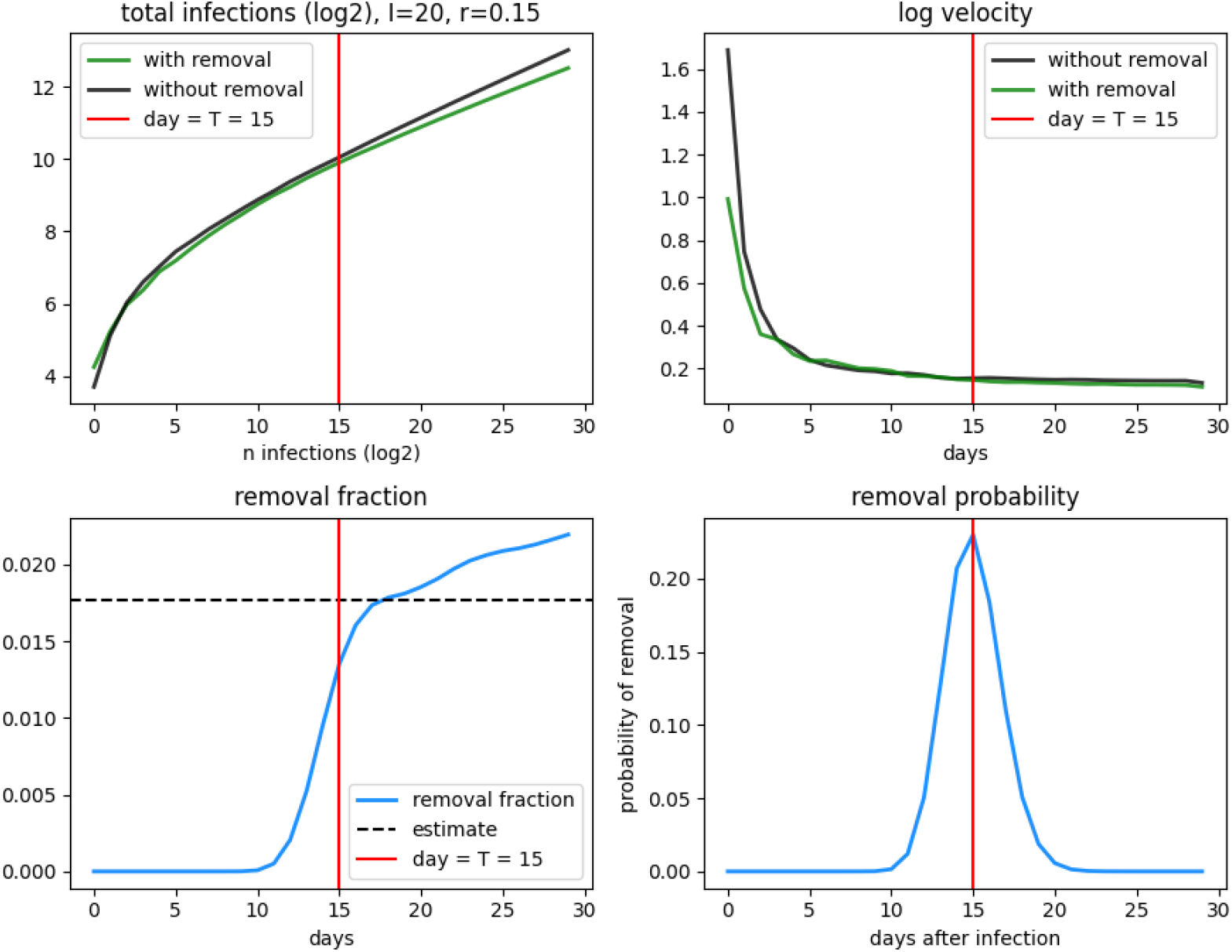
Drip model simulation with removal. We compare the dynamics for the drip model with and without removal. (upper left). The case counts for the two drip models with *r* = 0.15, *I* = 20 and *T* = 15. (upper right) The log-velocity of the two case counts considering removal (green) and without removal (black). (bottom left) The removal fraction (equation 36 computed numerically with the analytic estimate plotted in dashed black.) (bottom right) The removal probability used in the simulation: a discretized Gamma distribution with a mean of *T* = 15 days and a spread of 3 days.

**Table S1.** Symptoms grades for testing model.

| symptoms grade | description | symptoms |
| --- | --- | --- |
| 0 | asymptomatic | none |
| 1 | mild | common-cold like symptoms |
| 2 | severe | pneumonia-like symptoms, respiratory distress |

Note that the number of tests conducted on a given day is not, in general, equal to the number of *unique* tests conducted on a given day. This is because most countries implement protocols that call for duplicate testing. For now, we will not consider duplicate testing since we are only interested in the basic formulation and scaling of the testing dynamics.

## References

1. Lai MM, Cavanagh D. The molecular biology of coronaviruses. vol. 48. Elsevier; 1997.

2. Lu R, Zhao X, Li J, Niu P, Yang B, Wu H, et al. Genomic characterisation and epidemiology of 2019 novel coronavirus: implications for virus origins and receptor binding. The Lancet. 2020;395(10224):565–574.

3. Bai Y, Yao L, Wei T, Tian F, Jin DY, Chen L, et al. Presumed asymptomatic carrier transmission of COVID-19. Jama. 2020;323(14):1406–1407.

4. Guan Wj, Ni Zy, Hu Y, Liang Wh, Ou Cq, He Jx, et al. Clinical characteristics of coronavirus disease 2019 in China. New England journal of medicine. 2020;382(18):1708–1720.

5. Liu Y, Gayle AA, Wilder-Smith A, Rocklöv J. The reproductive number of COVID-19 is higher compared to SARS coronavirus. Journal of travel medicine. 2020;.

6. Li R, Pei S, Chen B, Song Y, Zhang T, Yang W, et al. Substantial undocumented infection facilitates the rapid dissemination of novel coronavirus (SARS-CoV-2). Science. 2020;368(6490):489–493.

7. World Health Organization Coronavirus disease 2019 (COVID-19): situation report, 72. 2020. Available from: https://www.who.int/emergencies/diseases/novel-coronavirus-2019/situation-reports

8. Tanne JH, Hayasaki E, Zastrow M, Pulla P, Smith P, Rada AG. Covid-19: how doctors and healthcare systems are tackling coronavirus worldwide. Bmj. 2020;368.

9. Lassaunière R, Frische A, Harboe ZB, Nielsen AC, Fomsgaard A, Krogfelt KA, et al. Evaluation of nine commercial SARS-CoV-2 immunoassays. medRxiv 2020.04.09.20056325 [Preprint] 2020 [cited 2014 May 10]. Available from : https://doi.org/10.1101/2020.04.09.20056325;

10. Organization WH, et al. Management of ill travellers at points of entry–international airports, seaports and ground crossings–in the context of COVID-19 outbreak: interim guidance, 16 February 2020. World Health Organization; 2020. Available from : https://apps.who.int/iris/bitstream/handle/10665/331003/WHO-2019-nCoV-POEmgmt-2020.1-eng.pdf

11. Tan J, Mu L, Huang J, Yu S, Chen B, Yin J. An initial investigation of the association between the SARS outbreak and weather: with the view of the environmental temperature and its variation. Journal of Epidemiology & Community Health. 2005;59(3):186–192.

12. Ortiz O, Castells F, Sonnemann G. Operational energy in the life cycle of residential dwellings: The experience of Spain and Colombia. Applied Energy. 2010;87(2):673–680.

13. Tono AMO, García M, Moncayo CJ, Wills C, Mahecha ÁMC. COVID-19: generalidades, comportamiento epidemiológico y medidas adoptadas en medio de la pandemia en Colombia. Acta de otorrinolarinología & cirugía de cabeza y cuello. 2020; p. 4–13.

14. Chen S, Chio C, Jou L, Liao C, et al. Viral kinetics and exhaled droplet size affect indoor transmission dynamics of influenza infection. Indoor Air. 2009;19(5):401.

15. Halloran SK, Wexler AS, Ristenpart WD. A comprehensive breath plume model for disease transmission via expiratory aerosols. PloS one. 2012;7(5).

16. Lin K, Marr LC. Humidity-dependent decay of viruses, but not bacteria, in aerosols and droplets follows disinfection kinetics. Environmental Science & Technology. 2019;54(2):1024–1032.

17. Kutter JS, Spronken MI, Fraaij PL, Fouchier RA, Herfst S. Transmission routes of respiratory viruses among humans. Current opinion in virology. 2018;28:142–151.

18. Yang W, Marr LC. Mechanisms by which ambient humidity may affect viruses in aerosols. Appl Environ Microbiol. 2012;78(19):6781–6788.

19. Herfst S, Böhringer M, Karo B, Lawrence P, Lewis NS, Mina MJ, et al. Drivers of airborne human-to-human pathogen transmission. Current opinion in virology. 2017;22:22–29.

20. Iwasaki A, Foxman EF, Molony RD. Early local immune defences in the respiratory tract. Nature Reviews Immunology. 2017;17(1):7.

21. Leung C. The difference in the incubation period of 2019 novel coronavirus (SARS-CoV-2) infection between travelers to Hubei and nontravelers: the need for a longer quarantine period. Infection Control & Hospital Epidemiology. 2020; p. 1–3.

22. Instituto Nactional de Salud Casos de COVID-19 en Colombia; 2020 [cited 2020 May 10] Database INS [Internet] Available from: https://www.ins.gov.co/Paginas/Boletines-casos-COVID-19-Colombia.aspx.

23. Ministerio de salud y protecctión social Lineamientos para el uso de pruebas diagnósticas de SARS-CoV-2 (COVID-19) en Colombia; 2020 [cited 2020 May 10] Database MSPS [Internet] https://www.minsalud.gov.co/Ministerio/Institucional/Procesos%20y%20procedimientos/GIPS21.pdf.

24. World weather online; 2020 [cited 2020 May 10] Database WWO [Internet] https://www.worldweatheronline.com/.

25. City Population; 2020 [cited 2020 May 10] Database [Internet] https://citypopulation.de/.

26. Aeronáutica Civil de Colombia Boletín Estadístico Enero 2019 Tráfico de Aeropuertos; 2019 [cited 2020 May 10] Database [Internet] http://www.aerocivil.gov.co/atencion/estadisticas-de-las-actividades-aeronauticas/_layouts/15/WopiFrame.aspx?sourcedoc=/atencion/estadisticas-de-las-actividades-aeronauticas/Estadsticas%20operacionales/Estadisticas%20Trafico%20de%20Aeropuertos%20Enero%202019.xls&action=default.

27. Moovit. Public Transit Index; 2020 [cited 2020 May 10] Database [Internet] https://moovitapp.com/insights/en/Moovit_Insights_Public_Transit_Index-countries.

28. Lowen A, Palese P. Transmission of influenza virus in temperate zones is predominantly by aerosol, in the tropics by contact: a hypothesis. PLoS currents. 2009;1.

29. Tang JW. The effect of environmental parameters on the survival of airborne infectious agents. Journal of the Royal Society Interface. 2009;6(suppl 6):S737–S746.

30. Chan K, Peiris J, Lam S, Poon L, Yuen K, Seto W. The effects of temperature and relative humidity on the viability of the SARS coronavirus. Advances in virology. 2011;2011.

31. Hemmes J, Winkler K, Kool S. Virus survival as a seasonal factor in influenza and poliomyelitis. Nature. 1960;188(4748):430–431.

32. Yang W, Elankumaran S, Marr LC. Relationship between humidity and influenza A viability in droplets and implications for influenza’s seasonality. PloS one. 2012;7(10).

33. Polozov IV, Bezrukov L, Gawrisch K, Zimmerberg J. Progressive ordering with decreasing temperature of the phospholipids of influenza virus. Nature chemical biology. 2008;4:248.

34. Brown JD, Goekjian G, Poulson R, Valeika S, Stallknecht DE. Avian influenza virus in water: infectivity is dependent on pH, salinity and temperature. Veterinary microbiology. 2009;136(1-2):20–26.

35. Chin AW, Chu JT, Perera MR, Hui KP, Yen HL, Chan MC, et al. Stability of SARS-CoV-2 in different environmental conditions. The Lancet Microbe. 2020;1(1):e10.

36. Prussin AJ, Schwake DO, Lin K, Gallagher DL, Buttling L, Marr LC. Survival of the enveloped virus Phi6 in droplets as a function of relative humidity, absolute humidity, and temperature. Appl Environ Microbiol. 2018;84(12):e00551–18.

37. Marr LC, Tang JW, Van Mullekom J, Lakdawala SS. Mechanistic insights into the effect of humidity on airborne influenza virus survival, transmission and incidence. Journal of the Royal Society Interface. 2019;16(150):20180298.

38. Walker JE, Wells RE. Heat and water exchange in the respiratory tract. The American journal of medicine. 1961;30(2):259–267.

39. Zhang L, Li Y. Dispersion of coughed droplets in a fully-occupied high-speed rail cabin. Building and Environment. 2012;47:58–66.

40. Page LA, Keshishian C, Leonardi G, Murray V, Rubin GJ, Wessely S. Frequency and predictors of mass psychogenic illness. Epidemiology. 2010;21(5):744–747.

41. Zaveri M. Sheriff Told Teen to Take Down Posts About Coronavirus, Family’s Lawsuit Says. The New York Times; April 21, 2020 [cited 2020 May 10].Available from : https://www.nytimes.com/2020/04/21/us/marquette-county-sheriff-instagram-lawsuit.html

42. Bhanot G, DeLisi C. Predictions for Europe for the Covid-19 pandemic from a SIR model. medRxiv. 10.1101/2020.05.26.20114058 [Preprint] 2020 [cited June 1 2020]. Available from https://doi.org/10.1101/2020.05.26.20114058

43. COVID-19: Data; The Official Website of the City of New York [Internet] https://www1.nyc.gov/site/doh/covid/covid-19-data.page.

